# Elevated levels of PM2.5 in crowded Subways of Cities with High COVID-19 related Mortality

**DOI:** 10.1101/2020.06.24.20138735

**Authors:** Yves Muscat Baron

## Abstract

**BACKGROUND:** Recent literature indicates that the pollutant, particulate matter PM2.5, may have an impact on COVID-19 related mortality. COVID-19 has been found adherent to PM2.5 and may be involved in the transmission and the exacerbation of COVID-19 infection, possibly due to PM2.5’s adverse influence on respiratory immunity. The PM2.5 levels in underground subways have been found up to 90 times higher than the surface levels. Moreover, the commuter congestion in the presence of such high levels of PM2.5 further encouraged COVID-19 human to human transmission.

**METHOD:** The levels of PM2.5 were retrieved from literature assessing particulate matter PM2.5 measured on subway platforms in two groups of cities. These cities were differentiated by the COVID-19 population percentage mortality rate (0.007% vs 0.09% (p<0.0004) the city’s population, more than a 10-fold difference. Data regarding the number of stations, length of the networks (km) and the annual ridership were also obtained from literature related to underground commuting.

**RESULTS:** The population percentage mortality related to COVID-19 infection correlated significantly for both minimum (p<0.01) and maximum (p<0.00001) levels of PM2.5. The cities’ subways with low COVID-19 mortality had minimum platform PM2.5 levels of 27.4 (SD+/-17.2µg/m3) compared to 63.4µg/m3 (SD+/-10.8µg/m3) in cities with high COVID-19 associated mortality (p<0.01). Subway maximum levels of PM2.5 in cities with low COVID-19 mortality was 53.4µg/m3 (SD+/- 21.8µg/m3) while that of underground networks with high COVID-19 mortality had maximum platform PM2.5 levels of 172.1µg/m3 (SD+/-98µg/m3) (p<0.001). The cities with higher COVID-19 mortality had longer networks 230km (SD+/-111km) versus 119km (SD+/-99km) (p<0.03) and more stations 191 (SD+/-109) versus 102 (SD+/-94) (p<0.047). Although the annual ridership in the cities with the high COVID-19 mortality was higher (1034×10^6^ vs 751×10^6^) this did not achieve statistical significance. The maximum PM2.5 correlated with the number of stations (p<0.045) and the length of the networks (p< 0.044). The minimum PM2.5 did not achieve similar significant correlations to subway variables. Ridership significantly correlated with number of stations (p<0.01) and the length of the network (p<0.02).

**CONCLUSION:** Underground networks may have inherent characteristics accelerating spread of COVID-19 infection and consequent mortality. The highly elevated levels of PM2.5 in overcrowded subways with extensive reach may have acted as a co-factor to disseminate the pandemic.

## INTRODUCTION

A nationwide study by Wu et al 2020 in the United States indicated a connection between long-term exposure to particulate matter PM2.5 and COVID-19 related mortality (Wu et al 2020). COVID-19 had been found to be adherent to particulate matter and it has been suggested that PM2.5 is an airborne co-factor in COVID-19 infection (Setti et al 2020). PM2.5 minimum and maximum levels in subways have been found several times higher than surface level. An elevated mean PM2.5 concentration of 381μg/m3 was reported on the Victoria Underground, approximately 16 times higher than surface PM2.5 results (Smith et al 2020). The World Health Organization guideline recommends that PM2.5 levels should not exceed an average level of 25μg/m^3^ 24-hour. The guideline stipulates that PM2.5 the concentration of 10μg/m^3^ is the lowest level at which combined cardiopulmonary and lung cancer mortality have been shown to rise in response to chronic exposure to PM2.5.

Commuter use of underground rail is significant representing around 11% of public transport journeys worldwide. In New York the daily ridership is 5 million and decreased by 90% after the institution of the New York lockdown (Harris 2020). More than 200 million passengers commute daily in the 156 underground networks around the world. With the movement of rural populations to urban areas as in China, the ridership is increasing as in subways of Beijing and Shanghai have recorded exponential increases.

## METHODOLOGY

The mortality rate from COVID-19 related deaths was obtained from 18 cities. Due to variable rates of COVID-19 testing and diagnoses between countries and cities, mortality as a marker for COVID-19 incidence was used as it is a more robust indicator of the occurrence of COVID-19 infection. Cities were selected as opposed to nations, because substantial regional differences in infection rates have noted within the same countries. One group of these cities did not have an elevated percentage of the population that succumbed to COVID-19 deaths. This group consisted, of Tokyo, Naples, Sydney Vienna, Hong Kong, Seoul, Toronto, Athens and Taipei. The second group had significantly higher COVID-19 mortality rates and included Tehran, Barcelona, Stockholm, Sao Paolo Wuhan, Paris, Milan, London and in New York. The mortality rates were obtained from various sources including the John Hopkins Corona Virus Resource Centre, the WHO website and other local sources.

The maximum and minimum levels particulate matter PM2.5, were obtained for both groups of city subways from the literature pertaining to PM2.5 measurement. An important caveat is that measurements between stations from the same underground networks due to the age of the subway, the depth and length of the subway and the mode of its ventilation. Variation to PM2.5 also occurs with the network ridership, relative humidity and seasonal changes may also occur.

Data regarding the underground networks of the city subways assessed were obtained from an internet site. The data retrieved included the number of stations, the length of the subway and the annual ridership for all networks.

## RESULTS

The percentage mortality of the individual’s cities’ population related to COVID-19 infection correlated significantly with the PM2.5 levels on the subway platforms. The correlation of the percentage population mortality with PM2.5 was significant for minimum PM2.5 levels (p<0.01) and highly significant for maximum (p<0.00001) levels of PM2.5 (Figure 1).

**Figure 1.**
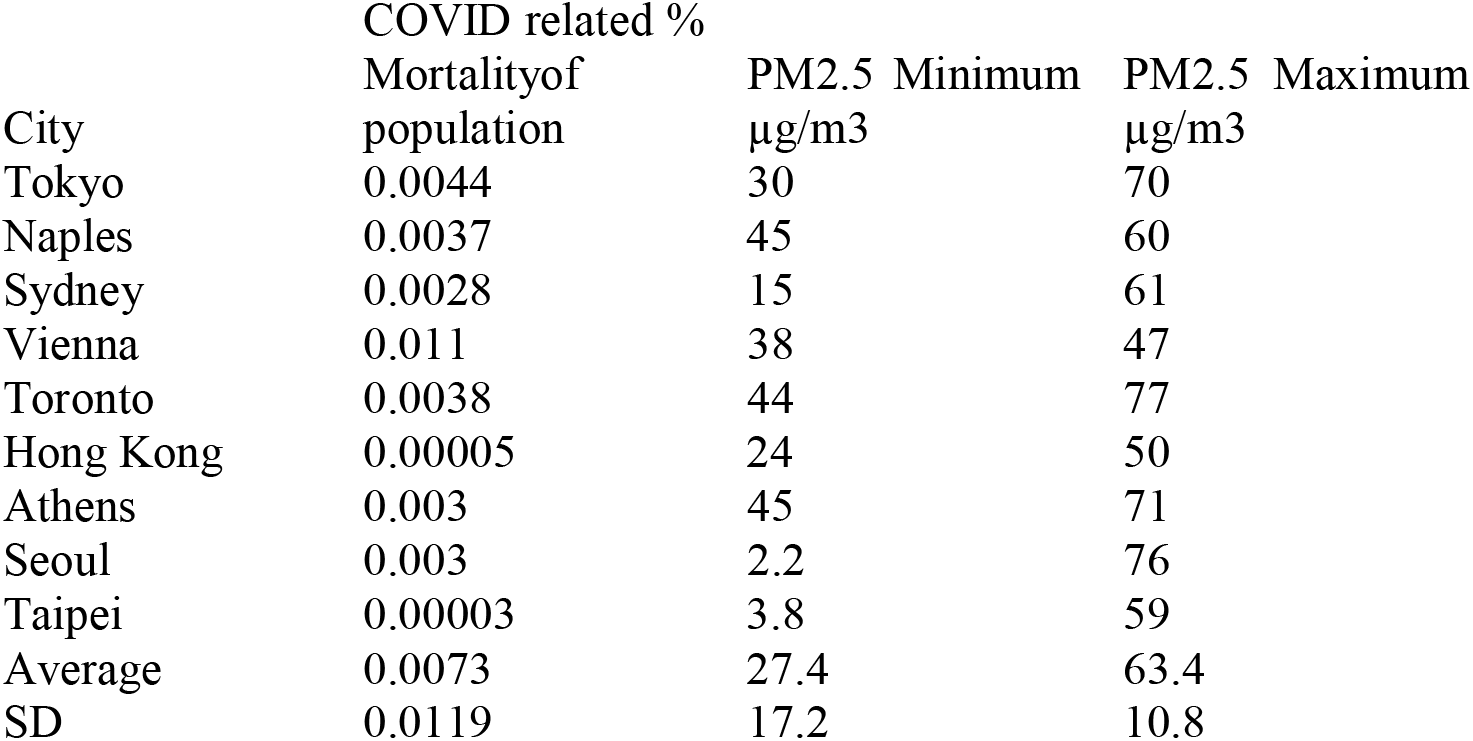
Low COVID related mortality as a percentage of the cities’ population and the associated minimum and maximum levels of PM2.5.

**Figure 2.**
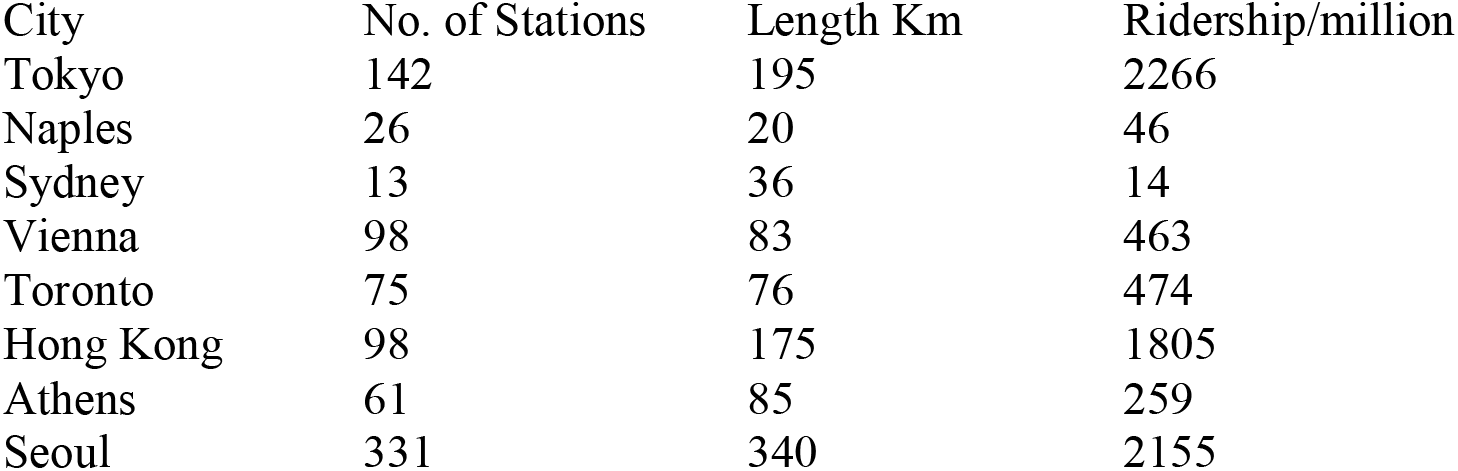
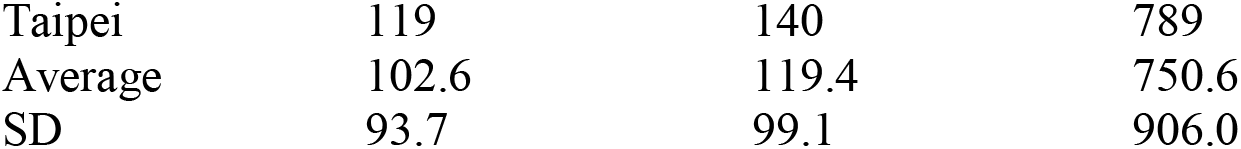
Cities with low COVID related mortality and characteristics of subway networks assessed.

The PM2.5 levels differed significantly between both groups of cities differentiated by COVID-19 related mortality. The cities’ subways with low COVID-19 death rates had minimum platform PM2.5 levels of 27.4 (SD+/-17.2 µg/m3) compared to 63.4µg/m3 (SD+/-10.8 µg/m3) in cities with high COVID-19 associated mortality rates (p<0.01). Underground networks’ maximum levels of PM2.5 in cities with low COVID-19 mortality was 53.4µg/m3 (SD+/-21.8µg/m3) while that of subways with high COVID-19 mortality had maximum platform PM2.5 levels of 172.1µg/m3 (SD+/-98µg/m3) (p<0.001) (Figures 1 and 3).

**Figure 3.**
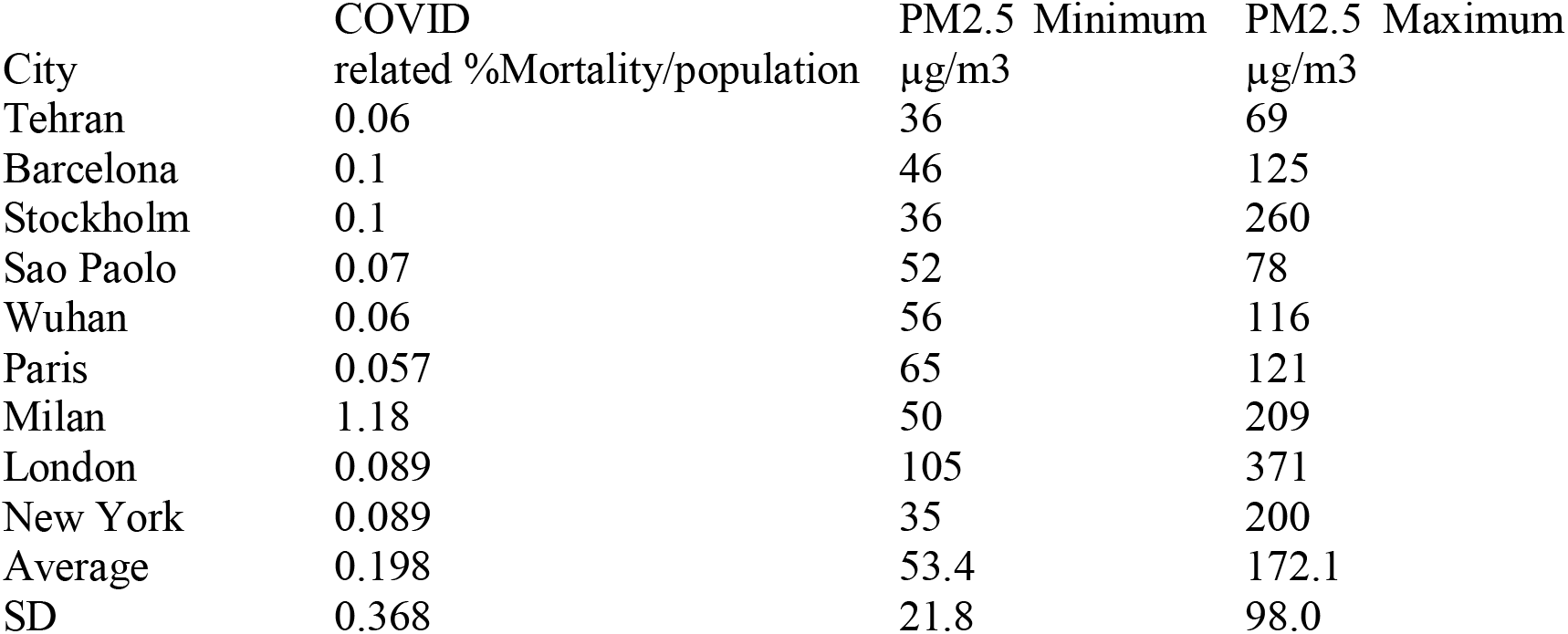
High COVID related mortality as a percentage of the cities’ population and the associated minimum and maximum levels of PM2.5.

The length of the subway network and number of stations was significantly different between both groups of cities. The cities with higher COVID-19 death rates had longer networks 230km (SD+/-111km) versus 119km (SD+/-99km) (p<0.03). Similarly cities with higher COVID-19 mortality rates had more stations 191 (SD+/- 109) versus 102 (SD+/-94), showing significance (p<0.047). Although the annual ridership in the cities with the high COVID-19 mortality was higher (1034×10^6^ versus 751×10^6^) this did not achieve statistical significance (Figures 2 and 4).

**Figure 4.**
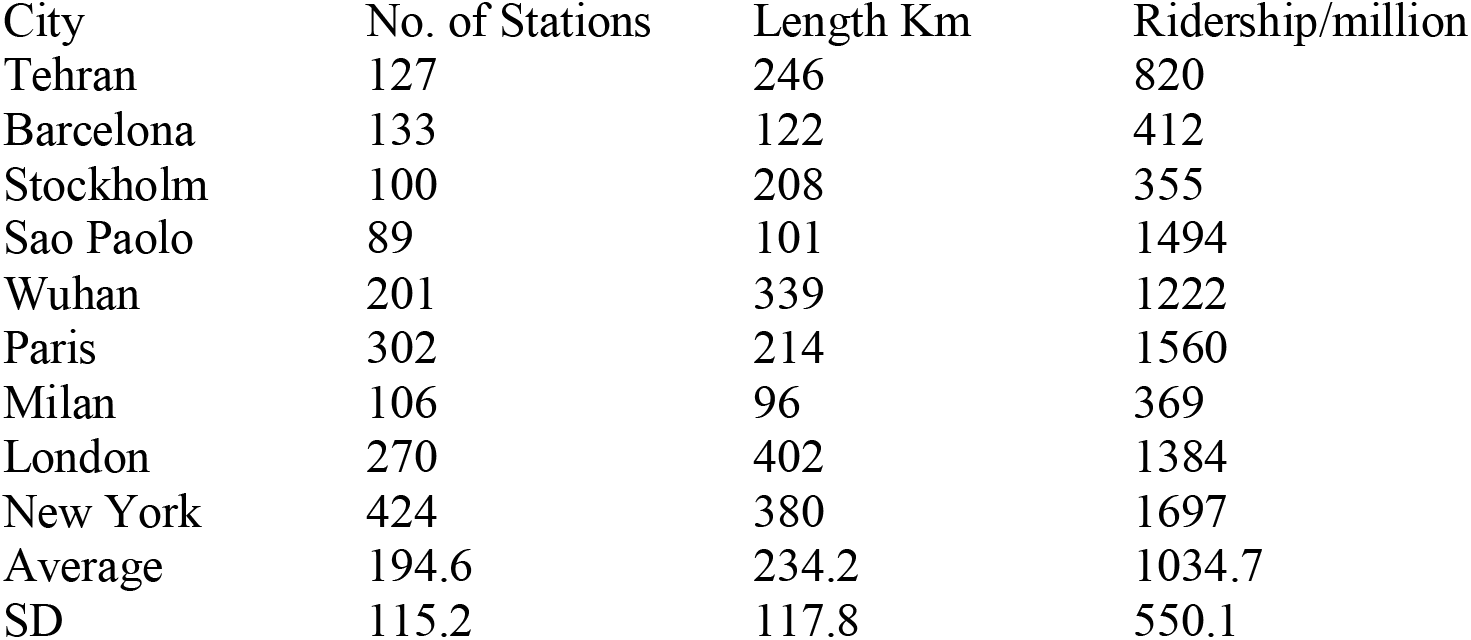
Cities with high COVID related mortality and characteristics of subway networks assessed.

The maximum PM2.5 correlated with the number of stations (p<0.045) and the length of the networks (p< 0.044). The minimum PM2.5 did not achieve similar significant correlations. Ridership significantly correlated with number of stations (p<0.01) and the length of the network (p<0.02).

## STATISTICS

The data was analysed for normality and all the data except for the minimum PM2.5 and the length of the networks were nonparametric. The Mann Whitney U test was applied for comparing nonparametric variables of both groups of cities and the Spearman Rank test was applied for nonparametric correlations.

## DISCUSSION

This review article indicated that the minimum and maximum levels of PM2.5 in underground networks correlated with the population percentage mortality rates associated with COVID-19 infection. The correlation between the population percentage mortality progressively increased from minimum (p<0.01) to maximum (p<0.00001) levels of PM2.5. This pattern of significance increases the biological plausibility of the correlation.

Underground stations are characterized by the congestion of numerous commuters confined to an enclosed space in the presence of very high levels the pollutant PM2.5. Elevated PM2.5 levels in overcrowded platforms and train cabins may have synergized possibly making the subways prime sites for seeding COVID-19.

PM2.5 concentrations in the London underground were found on average to be 18-times higher than concentrations at the surface. London underground levels of PM2.5 range from 39□μg□m−3 to 734□μg□m−3 on the deep tunnel lines and 14□μg□m−3 to 368□μg□m−3 for PM2.5 on the sub-surface lines (Saunders et al 2019). In another study the PM2.5 concentration in the London Underground (mean 88□μg□m−3, median 28□μg□m−3) was greater than at roadside environments (mean 22□μg□m−3, median 14□μg□m−3). PM2.5 mass varied up to a magnitude of 90 between lines and locations, with the subsurface and deepest lines being the District (median 4□μg□m−3) and Victoria (median 361□μg□m−3 but up to 885□μg□m−3) respectively (Smith et al 2019).

PM2.5 in underground networks is generated by several factors. These factors include friction between wheels and the rail and the braking mechanisms producing high levels of PM2.5. In this review the correlation between maximal PM2.5 with the number of stations supports the premise that braking mechanisms are responsible for high platform PM2.5 levels. Through the piston effect while trains travel through tunnels, the PM2.5 levels generated by wheel to rail friction would permeate the platform environment. This latter aspect would explain the PM2.5 correlation with the length of the subway networks.

High concentration of PM2.5 is more likely in confined areas as in subways as opposed to the surface. Dispersal by wind and rain do not occur in the underground. The temporary piston effect when trains pass through tunnels may disperse platform air but it may also push tunnel air towards the platform. Air quality of underground platform depends on a variety of factors including ventilation, train speed and frequency, wheel materials and braking mechanisms, and station depth and design (Moreno et al., 2014)

Several studies have shown adverse health impact associated with exposure to particulate matter PM2.5 (Brunekreef and Holgate, 2002, Pope and Dockery, 2006). Inhaled particulate matter has the propensity to create free radicals which at the pulmonary epithelium may cause oxidative stress, resulting in negative health effects (Kelly, 2003, Pourazar et al., 2005). The metallic components of subway PM2.5 have been shown to cause oxidative stress in alveolar macrophage cells (Steenhof et al., 2011, Jung et al., 2012).

The oxidative potential of particulate matter can be measured by assessing two antioxidants commonly found at the surface of the lung, ascorbic acid and glutathione. Ascorbic acid oxidation and glutathione oxidation shows low levels of oxidative potential possibly due to copper, arsenic and antimony emanating from brake pads and pantographs (Moreno et al 2016)

PM2.5’s proclivity to produce free radicals, peroxidation of cell membrane lipids occurs, with consequent rise of intracellular calcium. As a corollary elevated intracellular calcium increases inflammatory cytokine production (Kim et al 1997). PM2.5-induced inflammation led to an increase in the number of pulmonary neutrophils, eosinophils, T cells and mastocytes (Sigaud et al 2007)(Gripenbäck et 2005). All these cells can result in inflammatory cytokine production and resultant cytokine storm has been responsible for a significant number of COVID-19 related deaths (Nile et al 2020).

An experimental model has suggested that intervention on subway human traffic would have limited on the transmission of respiratory infection. (Cooley 2009). The model simulated the interactions of underground commuters with various sites of their daily activities. This model utilized information from the 1957–1958 H2N2 influenza epidemic and from New York City travel data. The model results indicated that the 1957-1958 pandemic had only a 4.4% contribution of influenza from subway interaction. This model concluded that public health interventions on the underground would have limited effect in containing the pandemic (Cooley 2009).

A crucial difference when comparing the 1957-58 influenza epidemic to COVID-19 is the reproduction number (R_0_) reflecting transmissibility of the viral infection. Whereas the R_0_ of the 1957-58 influenza epidemic was 1.1 at the beginning of 1957, the R_0_ of COVID-19 was estimated to be 3.28 in the initial stages (Liu et al 2020). This is substantially higher than the H2N2 in 1957 and also higher than severe acute respiratory syndrome (SARS) epidemic (R-factor 2.2) in 2003 (Liu et al 2020).

During the severe acute respiratory syndrome 2003 SARS epidemic, the air pollution index was noted to correlate with mortality. Patients who contracted SARS from Chinese provinces with high air pollution index had double the mortality compared to those from provinces with low air pollution index (RR =2.18, 95% CI: 1.31– 3.65). More specifically to particulate matter, PM10 and PM2.5 were implicated in the transmission of the Avian flu in 1997 (Chen et al 2010).

The above literature may suggest that the higher transmissibility of COVID-19 (R_0_ 3.28) may be due the presence of co-factors possibly including the utilization of particulate matter as a vector. COVID-19 genes have been found on particulate matter (Setti et al 2020). This study was carried out in the Bergamo region in Italy which was hard hit with COVID-19 mortality and also reports an elevated level of particulate matter pollution for both PM10 and PM2.5. As opposed to control samples of particulate matter, 34 RNA extractions for the COVID-19 E, N and RdRP genes, demonstrated 20 positive results for one of the genes (Setti et al 2020). This study suggested that airborne transmission was further augmented by ambient particulate matter. It is therefore biologically plausible that in the significantly higher concentrations of PM2.5 found in subways, the mode of transmission was further accelerated in the presence of overcrowded commuters on station platforms and train cabins.

This review indicated that subways in cities with high levels of COVID-19 mortality had significantly higher PM2.5 levels compared to cities with low deaths rates which concomitantly had low levels of PM2.5. The network characteristics such as the number of stations and rail track length correlated with PM2.5 concentration. The study by Wu et al showed that long-term exposure to PM2.5 increases COVID-19 related mortality. Just an increase of 1 μg/m3 in long-term PM2.5 exposure correlated with an 8% increase in the COVID-19 death rate. In a previous study by the same authors on a large American population of 65-year-olds, the same small increment in long-term exposure to PM2.5 led to an increase of 0.73% in all-cause mortality. Consequently for the same 1μg/m3 increase in PM2.5, the magnitude of COVID related deaths increased eleven-fold (Wu et al 2020).

The hypothesis of ambient salinity and the sodium chloride component in PM2.5 as a protective factor (Muscat Baron 2020a Muscat Baron 2020b) did not hold for the pandemic in the East Coast of USA. The states of New Jersey, Connecticut and Massachusetts have high levels of ambient salinity (Poma 2018). The role of subway interconnectivity and elevated PM2.5 levels may have eclipsed any protective factors causing high death rate in New York State (Harris 2020) and the adjacent states of New Jersey, Connecticut and Massachusetts. Due to the high connectivity New York, New Jersey and Connecticut are sometimes referred to as a Tristate.

This review has a number of limiting factors. Firstly although mortality related to COVID-19 infection is more robust that actual diagnosis of the infected cases. Mortality also depends in part on the testing prevalence and variance of diagnoses, especially in the presence of multiple pathology. This article did not go into depth as regards the date and the effect of lockdown on COVID-19 infection and mortality The implementation of national lockdown may have impacted the spread of infection however there was a spectrum of lockdowns throughout both groups of cities. Hong Kong and Seoul did not implement severely restrictive lockdowns. Both cities implemented tempestuous intervention to endorse mass testing and tracking. Moreover after the SARS epidemic the mindset of the people aided adhering to physical distancing in all its different facets including commuter use of subways (Park 2020).

Other limiting factors may also have affected the variables assessed. The PM2.5 levels in various cities were taken prior to the COVID-19 pandemic, during different seasons and years, and by different researchers and equipment. The characteristics of the subway networks may have varied prior to the pandemic and some are undergoing structural changes that may affect PM2.5 levels

## CONCLUSION

Overcrowded subways with elevated PM2.5 may expose commuters utilizing this mode of transport to higher viral loads adherent to particulate matter and consequent transmission. Combined with the adverse effects PM2.5 has on respiratory immunity, elevated subway PM2.5 levels in conjunction with inherent underground characteristics may possibly have increased COVID-19 infection spread and related mortality.

## Data Availability

Data are available from literature quoted in the extensive Reference section.

https://doi.org/10.1101/2020.04.05.20054502

https://doi10.3390/ijerph17082932

https://doi.org/10.1101/2020.05.03.20087056

https://doi10.21203/rs.3.rs-31884/v1

